# Accuracy, Clinical Utility and Usability Study of a Wireless Self-Guided Fetal Heart Rate Monitor

**DOI:** 10.1101/2020.11.18.20190959

**Authors:** Paul Porter, Fleur Muirhead, Joanna Brisbane, Brooke Schneider, Jennifer Choveaux, Natasha Bear, Jennie Carson, Kym Jones, Desiree Silva, Cliff Neppe

## Abstract

**Objective:** To evaluate the accuracy, reliability, clinical utility, and usability of HeraBEAT, a wireless fetal and maternal heart rate monitor (HBM) when used by clinicians and pregnant women to monitor fetal heart rate (FHR).

**Methods:** We recruited women aged 18 years or older with a singleton pregnancy of ≥12 weeks gestation. FHR recordings were performed using the HBM and cardiotocography (CTG) to determine comparative accuracy. The HBM was then used by clinicians and participants in the antenatal clinic with the latter then using the device unassisted to record at home. The women rated the HBM using the System Usability Scale (SUS).

**Results:** A total of 81 participants provided 126 recordings for analysis. The accuracy of the HBM was excellent compared with CTG, with limits of agreement (95%) between −1.5 and +0.9 beats per minute (bpm) and a mean difference of −0.29 bpm. The FHR was detected on 100% of occasions by clinicians (52 recordings) and participants when used in the clinic (42 recordings) and at home (32 recordings). Home users took an average of 1.1 minutes to detect the FHR and recorded a continuous trace of >1 minute in 94% of occasions, with an average total trace time of 4.4 minutes. The FHR trace was deemed to be clinically useful in 100% of clinician recordings and 97% of home recordings. There was no effect from body mass index, gestational age, pregnancy history, or placental position. The HBM ranked in the 96–100th percentile on the SUS for usability and learnability.

**Conclusions:** The HBM was accurate and easy for clinicians and participants to use. The data recorded at home was equivalent to that obtained in the clinic using current assessment protocols for low-risk pregnancies, allowing the device to be used in telehealth consultations.

**Clinical Trial Registration:** Australian New Zealand Clinical Trial Registry, https://www.anzctr.org.au ACTRN12620000739910.

## Introduction

The fetal heart rate (FHR) is an essential indicator of fetal well-being in utero. FHR monitoring is a standard component of antenatal and intrapartum care and is usually undertaken by clinicians using direct auscultation, handheld Doppler devices, or cardiotocography (CTG) machines.

In low-risk pregnancies, the FHR is often monitored for one minute in a process known as intermittent auscultation (IA). This is performed using a handheld Doppler, a DeLee-Hillis stethoscope, or a Pinard horn, depending on skills and resources. Although there is debate over the predictive value of IA for low-risk pregnancies, it is ubiquitously performed during routine antenatal examinations.^1–3^ In high-risk pregnancies, a nonstress test is performed using CTG to collect comprehensive FHR data for >10 minutes.^4^

Following recommendations by the Royal Australian and New Zealand College of Obstetricians and Gynaecologists to minimize patient contact during the COVID-19 pandemic, many clinical services have incorporated telehealth consultations into the antenatal program, replacing some face-to-face consultations. As we transition towards this new model, we need to examine how clinicians can deliver the same level of service. Many women already use electronic health (eHealth) technology and mobile applications (apps) to gather information and monitor their pregnancies. eHealth products are readily available and can engage and empower patients, resulting in benefits to health and well-being.^5–8^ However, whereas home monitoring of maternal parameters such as mindfulness, mood disorders, nausea, blood pressure, and weight^9–12^ is relatively straightforward, it has proven difficult to obtain clinically useful FHR traces at home.

Home FHR monitors are available, but there have been difficulties with usability, accuracy and reliability, signal noise, differentiation of fetal from maternal heart rate (MHR), inadequate recording duration, and cost.^13^ Handheld Doppler devices, which are used in clinics, require training to operate, cannot differentiate between FHR and MHR, and cannot store or transmit data. Although there have been attempts to use mobile CTGs, these machines are costly and not easily transportable. For home monitoring to be practical and clinically useful, FHR monitors need to be as accurate as CTGs, provide data that can be interpreted by clinicians, be self-administered, and allow secure and reliable data transmission. A home monitor should record a defined trace for at least 1 minute to determine whether the average FHR is within normal range (110–160 beats per minute [bpm]) and allow for evaluation of variability and accelerations.

HeraBEAT (HeraMED, Netanya, ISRAEL) is a medical-grade, low-cost, wireless, self-guided fetal and maternal heart-beat monitor (HBM) designed for self-administration from 12 weeks of gestation. It is held on the abdomen by the user, weighs 130 g, and is 9 cm in diameter (Figure 1). The HBM employs ultrawide beam Doppler technology and integrates a novel optical sensor to monitor the maternal heart rate directly from the abdomen, eliminating FHR-MHR cross talk. The HBM system includes a smartphone-based interface that guides device placement and displays the FHR trace and calculated parameters (average FHR and MHR using beat-to-beat calculation, duration of FHR trace, duration of search time, and longest continuous FHR segment) on a Bluetooth-connected smartphone. A printable recording of the fetal and maternal heart rates is produced for on-site or remote review (Figure 2). Clinicians can use a manual method to place the HBM directly on the appropriate position on the abdomen without voice guidance. The system is compliant with the HIPAA (Health Insurance Portability and Accountability Act) policies on privacy and transmission capabilities. The HBM system specifications and safety claims are presented in Appendix 1.

**Figure 1:**
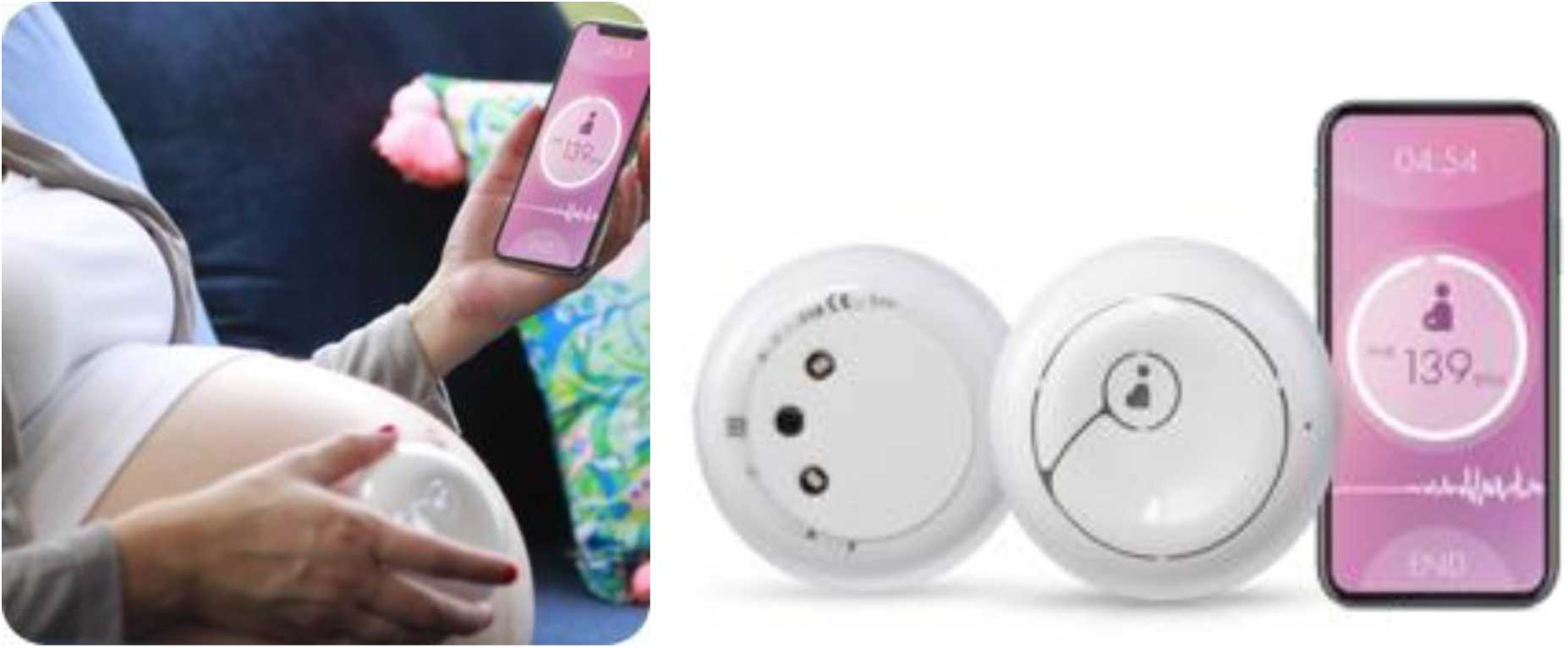
HeraBEAT system with device and integrated smartphone interface.

**Figure 2:**
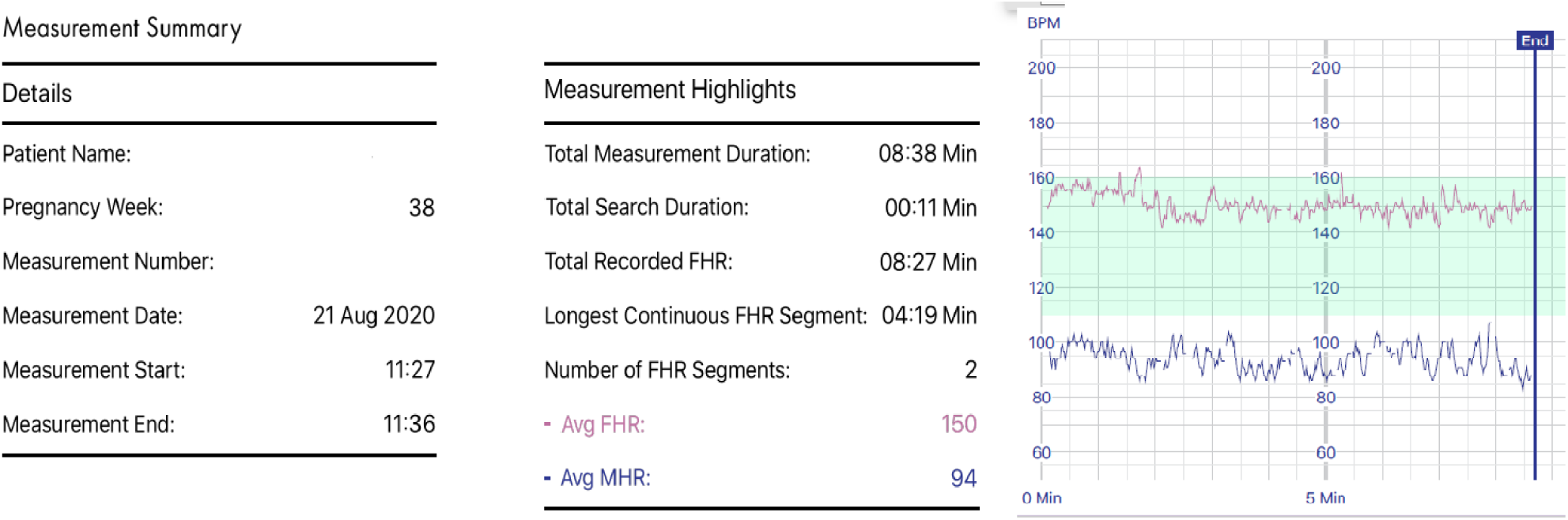
Data output from HeraBEAT system.

Our study objectives were to evaluate the accuracy, clinical utility, and operator usability of the HBM for both clinicians and pregnant women and to assess if the data generated were equivalent to those required by guidelines for IA.

## Methods

Ethical approval was obtained from the Ramsay Health Care Human Research Ethics Committee of Victoria and New South Wales (reference number 2020-005).

This was a prospective, single-center unblinded clinical study. We recruited participants as a convenience sample between July and September 2020 in the obstetrics department of a large metropolitan hospital in Western Australia. Potential participants were identified when presenting to the antenatal clinic. Women aged 18 years or older with a singleton pregnancy of at least 12 weeks gestation were approached to participate in the study. Women who were not able to read English, who had a skin rash or condition on the abdomen, or who had a pacemaker or other implantable electronic devices were excluded. Women who did not have access to a smartphone or internet connectivity were unable to participate in the home-recording sessions. Enrollment was undertaken by research nurses who explained the study and obtained written informed consent.

We collected data on women’s age, gestation, height, weight, body mass index (BMI), gravidity, parity, presence of a structural uterine abnormality, and location of the placenta, as well as data from the HBM.

Participants used the HBM in the self-guided mode at home, which uses the inbuilt position guidance system. The device is activated and placed below the umbilicus, as directed by the smartphone interface, to a position dependent on pregnancy gestation. The device continues to self-direct positioning using audio instructions until two distinct heart rates (FHR and MHR) are detected. Clinicians performed recordings in the antenatal clinic, where simultaneous monitoring using the HBM and an Avalon FM20 or Avalon FM30 CTG machine (Philips, Amsterdam, Netherlands) was undertaken to establish accuracy.

A research nurse showed participants how to use the HBM over a 5-minute training session and asked them to record data in the clinic and at home (self-monitoring). Participants were required to use the monitor unassisted to detect and record data for more than 1 minute. They were then asked to rate the HBM for usability and learnability. Home recordings were done between 1 and 21 days after clinic recordings.

We assessed the accuracy of the HBM compared with CTG by calculating paired FHR measurements taken at 15-second intervals for five sequential measures. Differences in FHR (bpm) between the paired measurements were analysed for each time point, for all time points combined, and for the mean of each subject’s five measurements. The agreement between HBM and CTG was established using Bland Altman plots and 95% limits of agreement. Furthermore 95% confidence intervals were established around the limits of agreement and displayed on the Bland Altman plots. Reliability was established using intraclass correlation coefficients using a two-way mixed effects model. The measurement comparison was deemed accurate if the 95% limits of agreement were within 8 bpm. This target was selected in keeping with other accuracy studies^14^ of FHR monitors as a clinically acceptable range in which important features, such as fetal bradycardias, accelerations, and decelerations, can be recognized.

From the recordings done by clinicians and from home recordings, we looked at the following outcome measures (1) detection of FHR (different from MHR), (2) number of continuous recordings longer than 1 minute, (3) total FHR recording time, (4) time taken to detect FHR, and (5) average FHR. We performed subgroup analyses to evaluate the relationship between BMI, gestation, obstetric history, and placental position on outcome measures for all participants and for the subgroup of women over 28 weeks gestation (in which CTG monitoring is typically performed). The relationship with clinical features was assessed using linear regression. When participants in the clinic used the HBM, the recordings were truncated at 1 minute and total trace times were not reported.

To examine clinical utility, obstetricians reviewed all recordings of over one minute to determine (1) if the FHR was in the normal range, (2) if separate FHRs and MHRs were detected, and (3) if FHR variability or accelerations were detectable during the duration of trace available. As all recordings were shorter than that required to establish a traditional baseline FHR (10 minutes), an average FHR (automatically calculated by the HBM) was used. The recordings from home were also assessed by obstetricians for quality of data after electronic transmission.

To assess the usability and learnability of the HBM, we used the international medical standard System Usability Scale (SUS).^HBM15^ The SUS is a 10-statement survey that evaluates the learnability, reliability, and usability of products. It has been shown to have high reliability (alpha of .91) over a wide range of interface types.^16^ When evaluating the results, SUS raw scores are reported as means and 95% confidence intervals and converted to a percentile rank (0-100) with a corresponding letter grade (A+ to F) as per the SUS Scoring system template (Appendix 2). Comparisons between clinic and home monitoring were performed using a paired t test. We used the positive version of the SUS and included an additional adjective rating scale, a single Likert scale question that demonstrates high correlation with overall SUS scores^17^ (Appendix 3). The adjectival rating scale the median and IQR range was provided given the skewed distribution Participants completed SUS questionnaires after using the HBM.

All data was analysed using Stata 14.1 (StataCorp, College Station, TX).

## Results

We enrolled 81 women into the study, and there were a total of 126 recordings available for analysis. The number of participants contributing data and the number of recordings obtained in each setting with each operator are shown in Figure 3. No adverse events were reported.

**Figure 3:**
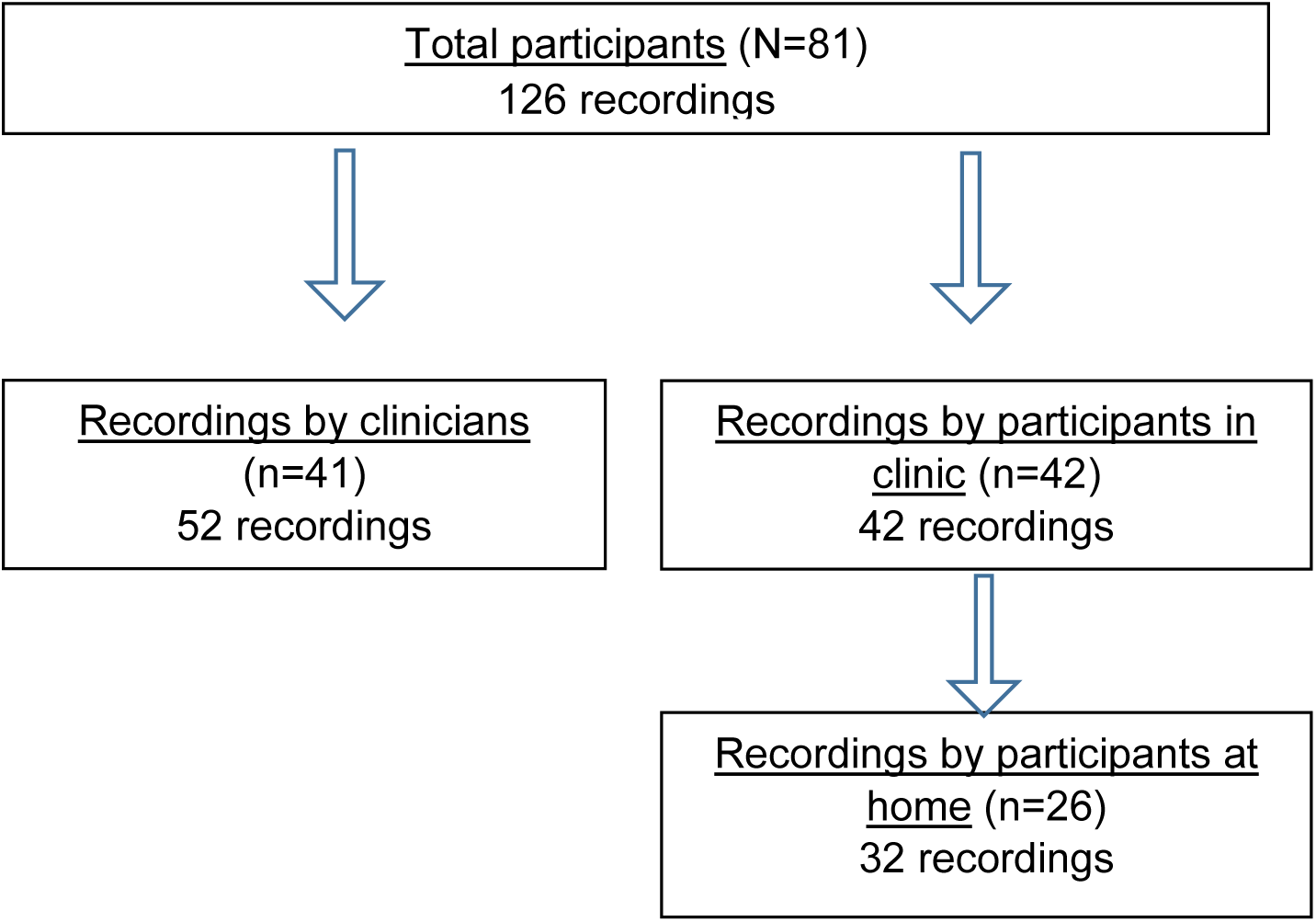
Patient involvement in study. Two subjects were included in both the clinician and participant arms.

Five participants contributed a recording in the clinic with a clinician and also self-recorded. Participants may have been involved in one or more recording sessions; however, all women who provided home recordings were trained to use the device in the clinic and also provided a recording in this setting. The clinical and demographic details of the study cohort are presented in Table 1.

**Table 1:** Demographic and clinical features.

| User | Clinician | Participant |  |
| --- | --- | --- | --- |
| Site of recording | Clinic (n=41) | Clinic (n=42) | Home (n=26) |
| Number of recordings | 52 | 42 | 32 |
| Age (y) | 30.5 ± 5.3 | 30.9 ± 4.2 | 31.7 ± 4.5 |
| Gestational age (wk) | 36.5 ± 3.5 | 31.1 ± 6.6 | 33.9 ± 5.6 |
| Pregnancy BMI (kg/m<sup>2</sup>) |  |  |  |
| <23.5 | 1 (2.4) | 4 (9.5) | 2 (7.7) |
| 23.5 to <30 | 18 (43.9) | 22 (52.4) | 10 (38.5) |
| 30 to <35 | 12 (29.3) | 11 (26.2) | 10 (38.5) |
| 35 to <45 | 10 (24.4) | 5 (11.9) | 4 (15.4) |
| 45+ | 0.0 (0.0) | 0 (0.0) | 0 (0.0) |
| Pregnancy BMI (kg/m<sup>2</sup>) |  |  |  |
| <35 | 31 (75.6) | 37 (88.1) | 22 (84.6) |
| ≥35 | 10 (24.4) | 5 (11.9) | 4 (15.4) |
| Gravidity |  |  |  |
| 1 | 16 (39.0) | 18 (42.9) | 14 (53.9) |
| 2 | 11 (26.8) | 14 (33.3) | 7 (26.9) |
| +3 | 14 (34.2) | 10 (23.8) | 5 (19.2) |
| Parity |  |  |  |
| 0 | 19 (46.4) | 19 (45.2) | 15 (57.7) |
| 1 | 11 (26.8) | 13 (31.0) | 7 (26.9) |
| +2 | 11 (26.8) | 10 (23.8) | 4 (15.4) |
| Placenta location |  |  |  |
| Posterior | 29 (70.7) | 30 (71.4) | 18 (69.2) |
| Anterior | 10 (24.4) | 6 (14.3) | 2 (7.7) |
| Lateral | 2 (4.9) | 4 (9.5) | 4 (15.4) |
| Fundal | 0 (0.0) | 2 (4.8) | 1 (3.9) |
| Low-lying | 0 (0.0) | 0 (0.0) | 1 (3.9) |
| Gestational age (wk) |  |  |  |
| 1st trimester (week 0–13) | 0 (0.0) | 2 (4.8) | 0 (0.0) |
| 2nd trimester (week 14–26) | 1 (2.4) | 7 (16.7) | 2 (6.3) |
| 3rd trimester (week 27+) | 51 (98.1) | 33 (78.6) | 30 (93.7) |

BMI, body mass index.
Data are mean ± SD or n (%).

We compared the accuracy of the HBM with CTG using simultaneous HBM and CTG recordings from 41 women. There were a total of 52 recordings and 260 paired data time points. Of the 260 paired measurements, the difference in FHR was ≤2 bpm for 249 (95.8%) paired measurements and between 3 and 5 bpm for 11 (4.2%) paired measurements. Characteristics of participants with a difference of >2 bpm between HBM and CTG at any given time point are shown in Appendix 4.

When all 260 time-paired data points were evaluated, the 95% limits of agreement between measurement devices were −2.982 bpm and 2.397 bpm, with a mean difference of −0.292 bpm (Figure 4). The intraclass correlation coefficient was 0.99.

**Figure 4:**
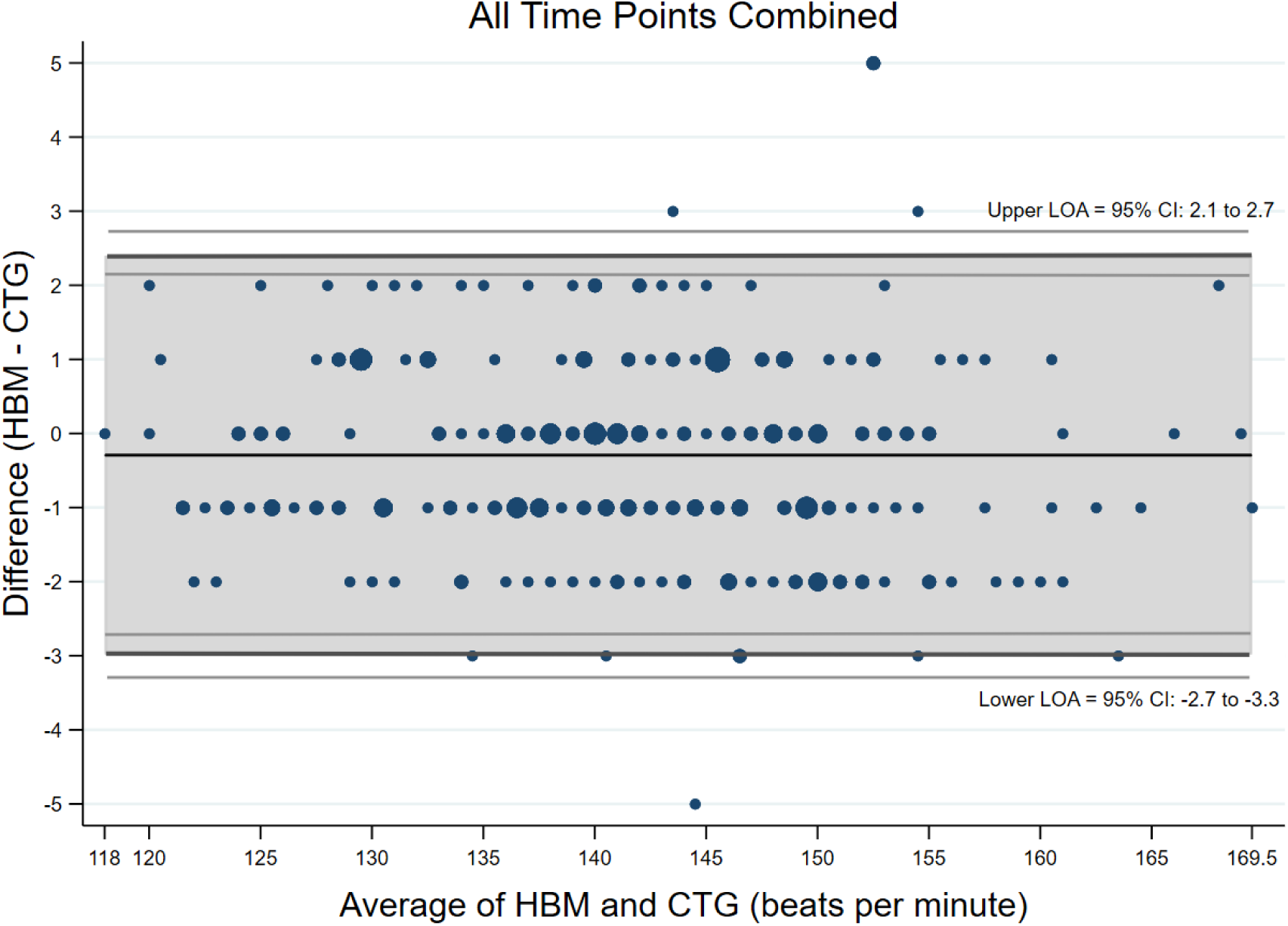
Bland-Altman plot showing comparable accuracy between HBM and CTG, with the difference in fetal heart rate (in beats per minute) between devices plotted across all time-paired data points (n=260). LOA, limits of agreement; CI, confidence interval; HBM, heartbeat monitor; CTG, cardiotocography.

When the difference between the means of the five time points for each device (n=52) was compared, the 95% limits of agreement were −1.478 bpm to 0.894 bpm, with a mean difference of −0.292 bpm (Figure 5). The intraclass coefficient was 0.99.

**Figure 5:**
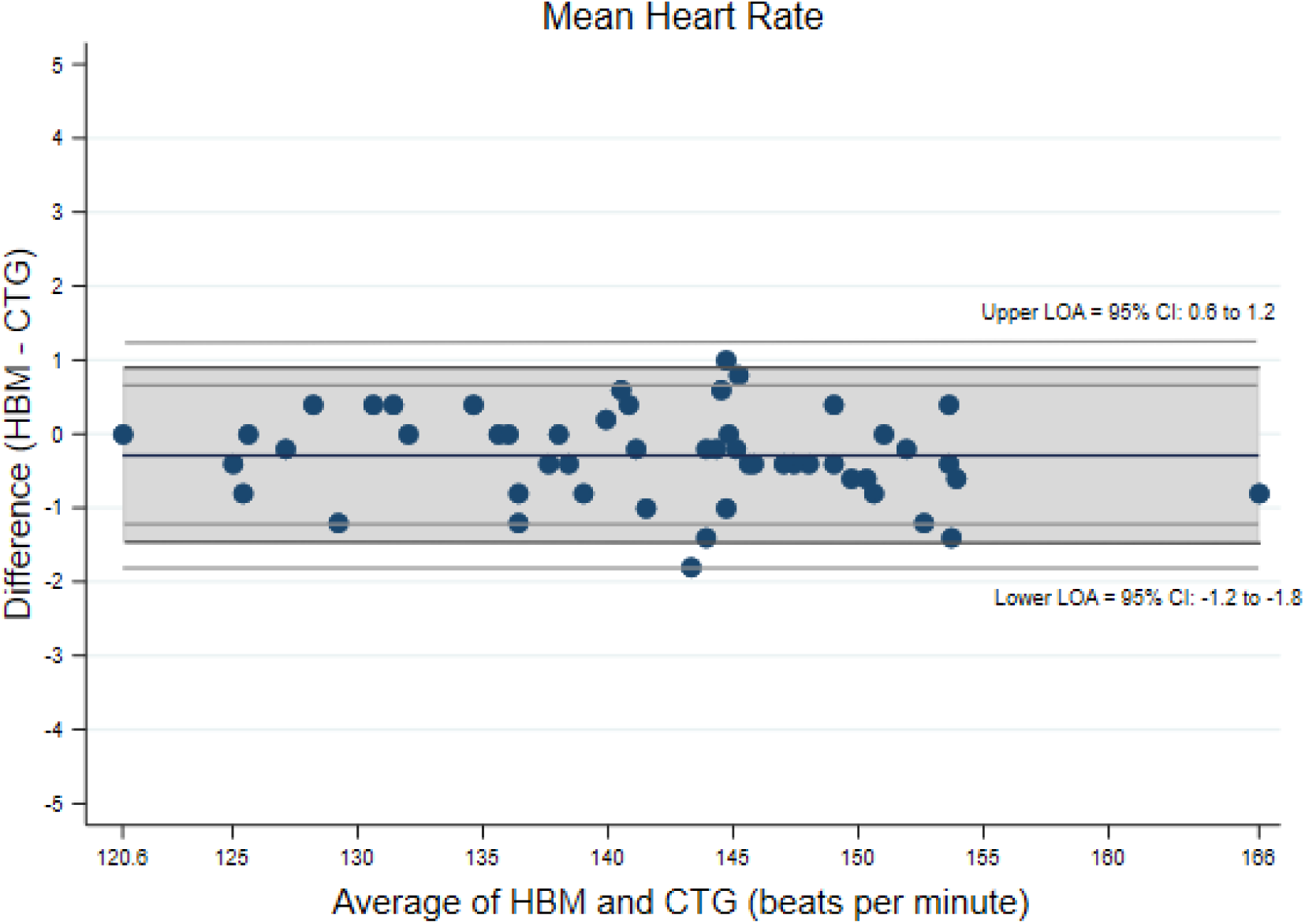
Bland-Altman plot showing comparable accuracy between HBM and CTG, with the difference in mean fetal heart rate between devices calculated over five time points (n=52). LOA, limits of agreement; CI, confidence interval; HBM, heart beat monitor; CTG, cardiotocography.

The FHR data obtained using the HBM are shown in Table 2. The FHR was detected in 100% of occasions by clinicians (52 recordings) and by 100% of participants (74 recordings) who used the device. The average time taken to detect a FHR was 0.9 minutes for clinicians (52 occasions), with 88.5% detected within 2 minutes; 0.7 minutes for self-recordings done in the clinic (42 occasions), with 92.8% detected within 2 minutes; and 1.1 minutes for home recordings (32 occasions), with 78% detected within 2 minutes.

**Table 2:** FHR metric data.

| FHR data | Clinician | Participant |  |
| --- | --- | --- | --- |
|  | Clinic<br>(n=41) | Clinic<br>(n=42) | Home<br>(n=26) |
| Number of recordings | 52 | 42 | 32 |
| Time to first detect a FHR (min) | 0.9 ± 0.9 | 0.7 ± 0.6 | 1.1 ± 1.7 |
| Average FHR (bpm) | 141.6 ± 9.6 | 141.5 ± 7.5 | 138.3 ± 9.8 |
| Average MHR (bpm) | 89.7 ± 9.4 | 83.2 ± 18.4 | NA |
| Continuous FHR trace duration (min) | 3.3 ± 2.2 | NA | 3.0 ± 1.3 |
| Total FHR trace duration (min) | 6.7 ± 1.9 | NA | 4.4 ± 0.7 |
| Tracings >1 min |  |  |  |
| Yes | 50 (96.2) | 37 (88.1) | 30 (93.7) |
| No | 2 (3.8) | 5 (11.9) | 2 (6.3) |
| Time to 1st detection of FHR (s) |  |  |  |
| <15 | 11 (21.2) | 3 (7.1) | 4 (12.5) |
| 15–<30 | 9 (17.3) | 12 (28.6) | 9 (28.1) |
| 30–<45 | 7 (13.5) | 14 (33.3) | 6 (18.8) |
| 45–<60 | 4 (7.7) | 2 (4.8) | 4 (12.5) |
| 60–<120 | 15 (28.8) | 8 (19.0) | 2 (6.2) |
| 120–<240 | 4 (7.7) | 2 (4.8) | 3 (9.4) |
| 240+ | 2 (3.8) | 1 (2.4) | 4 (12.5) |
| FHR detected |  |  |  |
| Yes | 52 (100.0) | 42 (100.0) | 32 (100.0) |
| No | (0.0) | (0.0) | (0.0) |
| MHR detected |  |  |  |
| Yes | 52 (100.0) | 42 (100.0) | NA |
| No | (0.0) | (0.0) | NA |
| Difference between FHR and MHR |  |  |  |
| Yes | 52 (100.0) | 42 (100.0) | NA |
| No | (0.0) | (0.0) | NA |
| Presence of variability |  |  |  |
| Yes | 52 (100.0) | NA | 31 (96.9) |
| No | (0.0) | NA | 1 (3.1) |
| FHR acceleration |  |  |  |
| Yes | 38 (73.1) | NA | 22 (62.5) |
| No | 14 (26.9) | NA | 10 (27.5) |

FHR, fetal heart rate; bpm, beats per minute; MHR, maternal heart rate; NA, not assessed. Data are mean ± SD or n (%).

A continuous FHR trace of >1 minute was recorded in 96.2% of occasions by clinicians, 88.1% of occasions by participants in the clinic, and by 93% of occasions by participants at home. The duration of the FHR trace was 6.7 minutes for clinicians and 4.4 minutes for home-monitoring participants.

To assess for clinical utility, obstetricians evaluated a total of 84 HBM traces, comprising 52 recordings from clinicians and 32 home recordings. A FHR was detected in 100% of occasions, and the traces were deemed to be clinically interpretable in 100% of clinician-obtained recordings and 31 of the 32 (96.9%) recordings by participants. One home-recorded trace was of insufficient duration to allow for evaluation of variability. FHR accelerations were present and identified in 73% of clinician recordings and 62.5% of home recordings (with average total trace durations of 6.7 minutes and 4.4 minutes, respectively). All home recordings were successfully transmitted to the clinical team with no data corruption.

Participants who used the HBM in the clinic (42 occasions) and at home (26 occasions) rated the usability and learnability of the HBM using the SUS. The mean total usability, reliability, and learnability scores ranked in the 96–100th percentile. The scores are presented in Table 3, together with their percentile and grades (A+ to F).^15,17^ There were no differences in SUS scores between clinic and home monitoring (*P*=.90, paired *t-test*). The adjectival rating scale, scored on a Likert scale of 1 to 7, gave a median score of 6 for both in-clinic and home use.

**Table 3:** System Usability Scale results for clinic and home use.

| Grading metric | Clinic use (n=42) |  |  | Home use (n=26) |  |  |
| --- | --- | --- | --- | --- | --- | --- |
|  | Total score | Usability | Learnability | Total score | Usability | Learnability |
| Raw SUS score | 89<br>(85.4–92.5) | 88.5<br>(85.2–92.3) | 89.9<br>(85.1–94.6) | 88.1<br>(82–94.2) | 86.9<br>(81.1–93.6) | 90.9<br>(84.9–96.8) |
| Percentile ranking | 96–100 | 96–100 | 96–100 | 96–100 | 96–100 | 96–100 |
| Graded score (A+ to F) | A+ | A+ | A+ | A+ | A+ | A+ |
| Adjectival rating scale (0–7) | 6 (6–7)<br>[5–7] |  |  | 6 (6–7)<br>[4–7] |  |  |

SUS, System Usability Scale.
Data are mean (95% confidence interval) or median (interquartile range) [range] unless otherwise specified.

There was no association between pregnancy variables including BMI, gestation, obstetric history, or recording site; and the time taken to detect a FHR, trace duration, or clinical utility of the HBM trace for the whole population and for pregnancies >28 weeks gestation. Two participants at 12 weeks gestation (2/2) successfully located the FHR and recorded continuous traces of > 1 minute. The time taken to detect FHR was shorter in the presence of an anterior placenta when a clinician used the HBM *(P=*.04) but not for participants. No other effect of placental position was seen. Data and *P* values for the effect of BMI, gestation, pregnancy history, and placental position are presented in Table 4. There were no differences when participants < 28 weeks gestation were excluded from the analysis.

**Table 4:** Factors related to clinical outcomes.

| Outcome | Clinician |  | Participant |  |  |  |
| --- | --- | --- | --- | --- | --- | --- |
|  | Clinic |  | Clinic |  | Home |  |
|  | Mean (SD) | P | Mean (SD) | P | Mean (SD) | P |
| Time to 1st detection of FHR (s) |  |  |  |  |  |  |
| Pregnancy BMI (kg/m<sup>2</sup>) |  |  |  |  |  |  |
| <23.5 | 0.3 (0.0) | .74 | 0.7 (0.6) | .77 | 0.6 (0.2) | .76 |
| 23.5 to <30 | 1.0 (1.1) |  | 0.6 (0.5) |  | 0.9 (1.0) |  |
| 30 to <35 | 0.7 (0.6) |  | 0.9 (0.9) |  | 1.2 (2.5) |  |
| 35 to <45 | 0.9 (0.6) |  | 0.6 (0.4) |  | 1.8 (1.5) |  |
| 45+ | 0 (0.0) |  | 0 (0.0) |  | 0 (0.0) |  |
| BMI ≤35 (kg/m<sup>2</sup>) |  |  |  |  |  |  |
| <35 | 0.9 (0.9) | .88 | 0.7 (0.7) | .82 | 1.0 (1.7) | .34 |
| ≥35 | 0.9 (0.6) |  | 0.6 (0.4) |  | 1.8 (1.5) |  |
| Anterior placenta location |  |  |  |  |  |  |
| Yes | 0.4 (0.4) | .04 | 0.6 (0.5) | .08 | 2.0 (1.4) | .46 |
| No | 1.0 (0.9) |  | 1.1 (1.0) |  | 1.1 (1.7) |  |
| Gestation |  |  |  |  |  |  |
| 1st trimester (week 0–13) | 0 (0.0) | .66 | 0.5 (0.0) | .17 | 0 (0.0) | .75 |
| 2nd trimester (week 14–26) | 1.3 (0.0) |  | 1.1 (0.7) |  | 0.8 (0.4) |  |
| 3rd trimester (week 27+) | 0.9 (0.0) |  | 0.6 (0.6) |  | 1.2 (1.7) |  |
| Continuous FHR trace duration (min) |  |  |  |  |  |  |
| Pregnancy BMI |  |  | NA | NA |  |  |
| <23.5 | 3.3 (0.0) | .81 |  |  | 2.2 (0.6) | .23 |
| 23.5 to <30 | 3.5 (2.5) |  |  |  | 3.4 (1.3) |  |
| 30 to <35 | 2.8 (1.5) |  |  |  | 3.1 (1.3) |  |
| 35 to <45 | 3.2 (2.1) |  |  |  | 2.1 (1.1) |  |
| 45+ | 0.0 (0.0) |  |  |  | 0.0 (0.0) |  |
| BMI ≤35 (kg/m<sup>2</sup>) |  |  | NA | NA |  |  |
| <35 | 3.3 (2.2) | .86 |  |  | 3.2 (1.3) | .10 |
| ≥35 | 3.2 (1.9) |  |  |  | 2.1 (1.1) |  |
| Anterior placenta location |  |  | NA | NA |  |  |
| Yes | 3.0 (1.8) | .61 |  |  | 2.4 (1.6) | .51 |
| No | 3.4 (2.3) |  |  |  | 3.0 (1.3) |  |
| Gestation |  |  | NA | NA |  |  |
| 1st trimester (week 0–13) | 0 (0.0) | .90 |  |  | 0 (0.0) | .94 |
| 2nd trimester (week 14–26) | 3.0 (0.0) |  |  |  | 2.9 (0.9) |  |
| 3rd trimester (week 27+) | 3.3 (2.2) |  |  |  | 3.0 (1.3) |  |

## Discussion

The results of this study show that the HBM is accurate and easy to use for clinicians in the hospital and for pregnant women at home. The FHR data obtained at home is equivalent to that required in the clinic using current IA protocols for low-risk pregnancies, which could allow for the device to be used during telehealth consultations.

The 95% limits of agreement between mean FHRs from the HBM and CTG were −1.5 and 0.9 bpm (mean difference −0.3bpm) which is well within the acceptability limit of 8 bpm. The limits of agreement were maintained across the FHR range (118-170bpm), BMI, gestational age and placental position. A FHR was detected in 100% of clinician-recordings (n=52) and 100% of self-recordings (42 from the clinic; 32 from home). FHR traces were clinically interpretable in 100% of clinician-performed and in 97% of participant-administered recordings. The HBM was ranked highly (96–100% percentile) for usability, reliability, and user satisfaction.

Based on these results, the HBM could facilitate the evolution of antenatal care models to incorporate telehealth consultations and remote self-monitoring. For clinicians, the monitor may be used as an alternative to handheld Doppler devices for IA in antepartum and intrapartum care.

Although most protocols recommend an initial face-to-face consultation by 10 weeks gestation and a total of 12-14 visits for uncomplicated pregnancies,^18^ the evidence to support this is not robust. Studies have suggested that reducing this number is safe,^19,20^ but there has been little change. The resistance may relate to concerns over adequate fetal monitoring and a perception that obstetricians’ engagement is tied to personal interactions.^21,22^ Several groups have tried to implement remote models by substituting face-to-face with telehealth consultations. The move away from a sickness care model has been associated with improved confidence, satisfaction, and empowerment for women.^21^ In addition to immediate health gains, the use of targeted technological aids has direct cost-saving advantages. The cost of US prenatal care may be reduced by 2.5% to 13% by using digital tools during telehealth consultations.^20^

During the COVID-19 pandemic, the Royal Australian and New Zealand College of Obstetrics and Gynaecology (along with other regulatory bodies) have recommended changes including reducing, postponing, or increasing the interval between face-to-face visits; limiting consultations to under 15 minutes and using telehealth consultations as an alternative; and cancelling in-person antenatal classes.^23^ For telehealth programs to be effective, there must be robust surveillance of maternal and fetal biomarkers. The accuracy and usability of the HBM, as well as its ability to store and transmit data, positions it strongly as a device that can be used in these programs.

IA is routinely performed to monitor FHR despite its uncertain predictive value. Studies have shown that regular FHR monitoring is associated with improved maternal well-being, satisfaction, and engagement.^24^ Detection of a FHR in the healthy range is the first aim of IA, but recognition of variability is dependent on clinical experience and accelerations are only noted if they occur within the 1-minute window. This short period of monitoring is not usually stored for review, and is heavily reliant on clinician experience. The HBM has advantages over handheld Doppler devices for IA, including cost, ability to distinguish maternal and fetal heart rates, ease of use for self-administration, data storage, and transmission capabilities.

Globally, as many as 2 million babies die during labor each year.^9,11,12,25^ The International Federation of Gynecology and Obstetrics guidelines recommend IA for 1 minute during antenatal care and labor when there is no access to CTGs.^26^ In resource-limited settings, it is common for this to be done by direct auscultation, although handheld Doppler devices are preferred because of their accuracy, readable displays, and comfort. In these settings, the HBM would allow even inexperienced operators to record, store, and transmit FHR data accurately. IA is also appropriate and recommended for intrapartum monitoring in low-risk pregnancies, including home births.^26^ The HBM could also be used in this setting, but further research is required to see if the device remains accurate during contractions.

Our study has several limitations, including the duration of tracings. Based on requirements for IA in antenatal and intrapartum settings, we focused on recording FHRs for at least 1 minute. Our study times significantly exceeded this, but we did cap home recordings to 5 minutes and our findings are not equivalent to nonstress test examinations. In high-risk pregnancies and labor, a 10- to 20-minute CTG recording is recommended to assess baseline FHR, variations, accelerations, and decelerations. Our HBM traces were too short to allow evaluation for decelerations and were not collected during contractions. We used an automatically generated average FHR based on beat-to-beat measurements and then assessed variability and accelerations from that baseline. Despite the short recordings, accelerations were evident in 73% of cases.

Our population was recruited from a single center and excluded women who could not read English and those with no access to a smartphone. However, as positional guidance is provided verbally by the smartphone interface and is available in multiple languages, we expect that language would not be an obstacle to broader use.

In this study, home recordings were collected in the self-guiding mode, and the time taken to detect a FHR was around 1 minute. The device also has a manual mode, which allows for experienced users to position the device without guidance (similar to Doppler devices). This may result in quicker detection of FHR by clinicians and would make for an interesting comparison study.

A strength of this study was the ability to compare HBM with CTG data, the latter being the gold standard for FHR monitoring. Additionally, we had a diverse study population in terms of gestation (12–40 weeks), BMI, placental positions, and obstetric history. Data accuracy, quality, and usability were maintained across these groups. There were no negative effects from higher BMIs or anterior placentas, conditions that could theoretically interfere with ultrasound detection of FHR. We selected a robust learnability and usability rating system which is widely used to evaluate medical devices.

We have shown that women are able to use the HBM at home to perform accurate and clinically relevant monitoring of FHR. The device addresses a critical hurdle for telehealth consultations and may offer confidence in the transition towards this new model of service.

## Data Availability

Unidentified data is available from the corresponding author for analysis dependent upon obtaining reciprocal ethical approval for sharing.

## Supplementary Files

## Appendix 1

HeraBEAT system specification and safety claims.

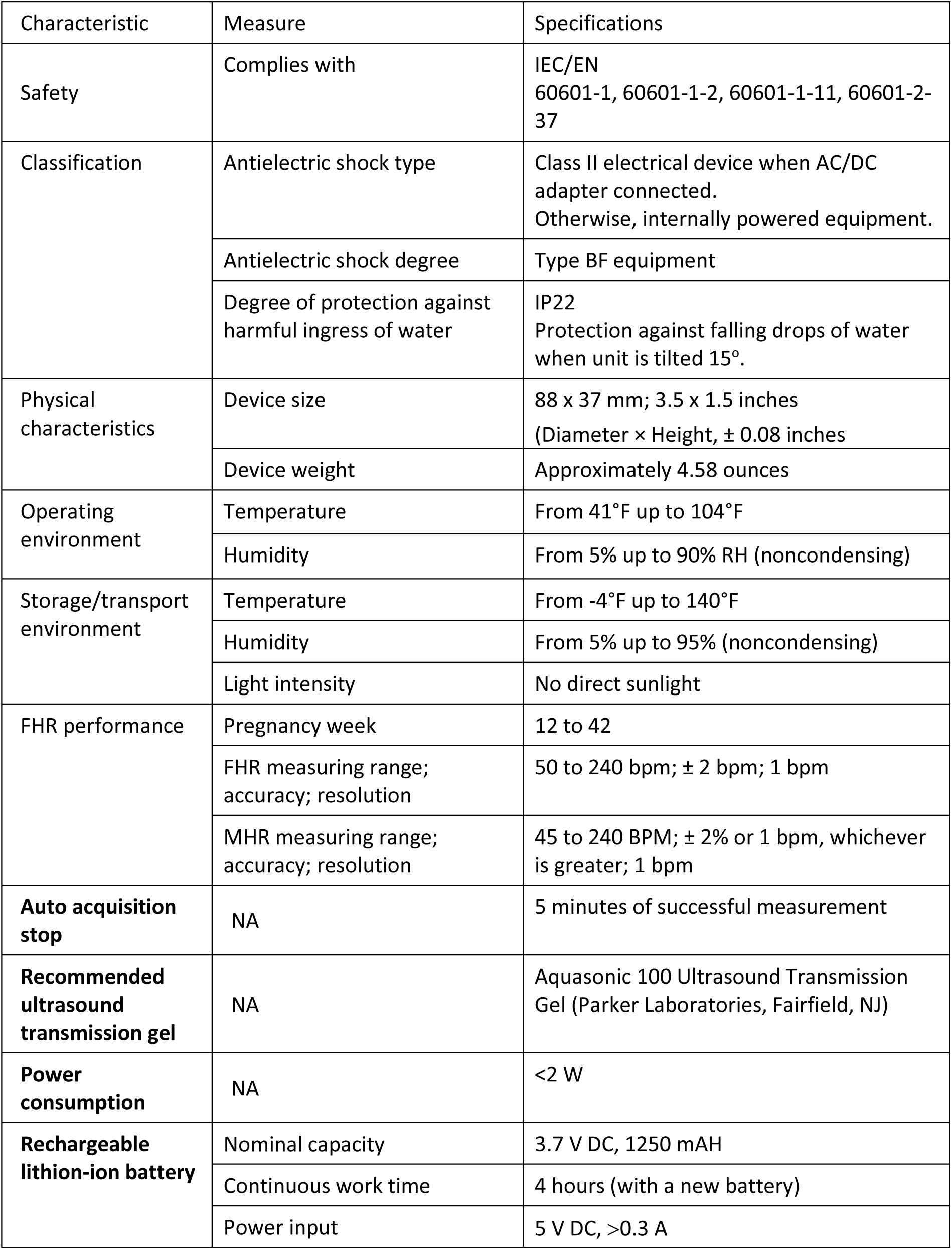
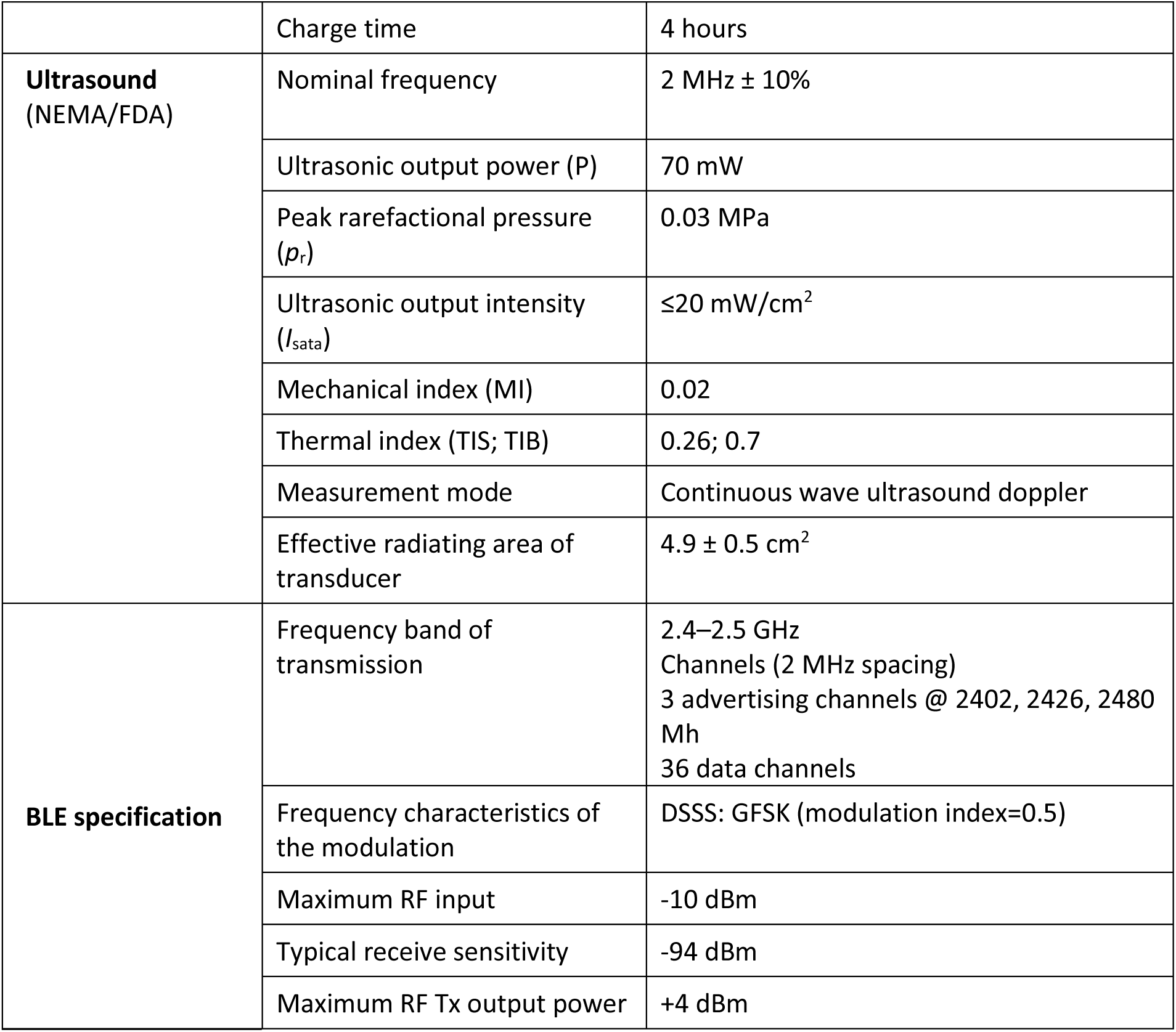

| Characteristic | Measure | Specifications |
| --- | --- | --- |
| Safety | Complies with | IEC/EN 60601-1, 60601-1-2, 60601-1-11, 60601-2-37 |
| Classification | Antielectric shock type | Class II electrical device when AC/DC adapter connected.<br>Otherwise, internally powered equipment. |
|  | Antielectric shock degree | Type BF equipment |
|  | Degree of protection against harmful ingress of water | IP22<br>Protection against falling drops of water when unit is tilted 15°. |
| Physical characteristics | Device size | 88 x 37 mm; 3.5 x 1.5 inches<br>(Diameter × Height, ± 0.08 inches) |
|  | Device weight | Approximately 4.58 ounces |
| Operating environment | Temperature | From 41°F up to 104°F |
|  | Humidity | From 5% up to 90% RH (noncondensing) |
| Storage/transport environment | Temperature | From -4°F up to 140°F |
|  | Humidity | From 5% up to 95% (noncondensing) |
|  | Light intensity | No direct sunlight |
| FHR performance | Pregnancy week | 12 to 42 |
|  | FHR measuring range; accuracy; resolution | 50 to 240 bpm; ± 2 bpm; 1 bpm |
|  | MHR measuring range; accuracy; resolution | 45 to 240 BPM; ± 2% or 1 bpm, whichever is greater; 1 bpm |
| **Auto acquisition stop** | NA | 5 minutes of successful measurement |
| **Recommended ultrasound transmission gel** | NA | Aquasonic 100 Ultrasound Transmission Gel (Parker Laboratories, Fairfield, NJ) |
| **Power consumption** | NA | <2 W |
| **Rechargeable lithium-ion battery** | Nominal capacity | 3.7 V DC, 1250 mAh |
|  | Continuous work time | 4 hours (with a new battery) |
|  | Power input | 5 V DC, >0.3 A |
|  | Charge time | 4 hours |
| **Ultrasound**<br>(NEMA/FDA) | Nominal frequency | 2 MHz ± 10% |
|  | Ultrasonic output power (P) | 70 mW |
| | Peak rarefactional pressure ($p_r$) | 0.03 MPa |
| | Ultrasonic output intensity ($I_{sata}$) | $\leq 20$ mW/cm<sup>2</sup> |
|  | Mechanical index (MI) | 0.02 |
|  | Thermal index (TIS; TIB) | 0.26; 0.7 |
|  | Measurement mode | Continuous wave ultrasound doppler |
| | Effective radiating area of transducer | 4.9 ± 0.5 cm<sup>2</sup> |
| **BLE specification** | Frequency band of transmission | 2.4–2.5 GHz<br>Channels (2 MHz spacing)<br>3 advertising channels @ 2402, 2426, 2480 Mh<br>36 data channels |
|  | Frequency characteristics of the modulation | DSSS: GFSK (modulation index=0.5) |
|  | Maximum RF input | -10 dBm |
|  | Typical receive sensitivity | -94 dBm |
|  | Maximum RF Tx output power | +4 dBm |

HeraBEAT safety claims:

- HeraBEAT works at low voltage (5 V), which is supplied from an internal rechargeable battery (tested per IEC 60601-1).
- HeraBEAT device material is isolated and made of electric nonconducting material. In addition, the device does not operate while charging.
- HeraBEAT transmits ultrasonic energy at a maximum intensity of 20 mW/cm^2^, according to IEC 60601-2-37 “Medical electrical equipment – Part 2-37: Particular requirements for the safety of ultrasonic medical diagnostic and monitoring equipment.”
- The device turns off if not connected to the mobile app for several seconds.
- All materials are biocompatible and approved for use on the skin surface.
- HeraBEAT controls the temperature level inside the device to assure that the device temperature remains below the safe temperature limit. In addition, a built-in test (BIT) is implemented to verify the correct functioning of the temperature sensor.
- The device conforms to risk management best practices according to ISO 14971:2007 – Medical Devices – Application of Risk Management to Medical Devices.

## Appendix 2

Conversion table for System Usability Scale raw scores into percentile and grades.

| SUS Score | Percentile | Grade |
| --- | --- | --- |
| 84.1–100 | 96–100 | A+ |
| 80.8–84.0 | 90–95 | A |
| 78.9–80.7 | 85–89 | A- |
| 77.2–78.8 | 80–84 | B+ |
| 74.1–77.1 | 70–79 | B |
| 72.6–74.0 | 65–69 | B- |
| 71.1–72.5 | 60–64 | C+ |
| 65.0–71.0 | 41–59 | C |
| 62.7–64.9 | 35–40 | C- |
| 51.7–62.6 | 15–34 | D |
| <51.7 | 0–14 | F |

## Appendix 3

All positive version of the System Usability Scale and the adjectival enhancement question used in the study.

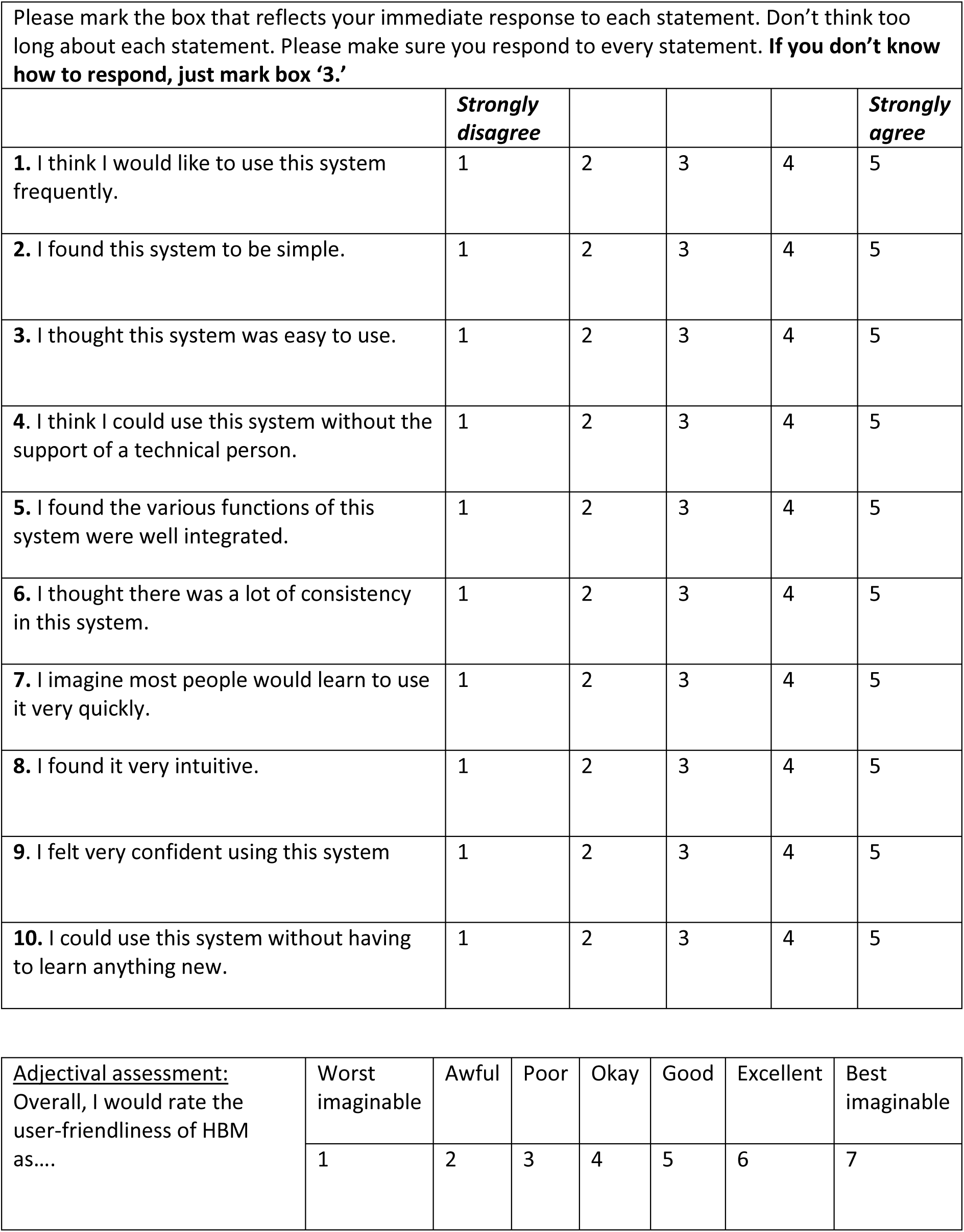

|  |  |  |  |  |  |
| --- | --- | --- | --- | --- | --- |
| Please mark the box that reflects your immediate response to each statement. Don't think too long about each statement. Please make sure you respond to every statement. **If you don't know how to respond, just mark box '3.'** |  |  |  |  |  |
|  | **Strongly disagree** |  |  |  | **Strongly agree** |
| 1. I think I would like to use this system frequently. | 1 | 2 | 3 | 4 | 5 |
| 2. I found this system to be simple. | 1 | 2 | 3 | 4 | 5 |
| 3. I thought this system was easy to use. | 1 | 2 | 3 | 4 | 5 |
| 4. I think I could use this system without the support of a technical person. | 1 | 2 | 3 | 4 | 5 |
| 5. I found the various functions of this system were well integrated. | 1 | 2 | 3 | 4 | 5 |
| 6. I thought there was a lot of consistency in this system. | 1 | 2 | 3 | 4 | 5 |
| 7. I imagine most people would learn to use it very quickly. | 1 | 2 | 3 | 4 | 5 |
| 8. I found it very intuitive. | 1 | 2 | 3 | 4 | 5 |
| 9. I felt very confident using this system | 1 | 2 | 3 | 4 | 5 |
| 10. I could use this system without having to learn anything new. | 1 | 2 | 3 | 4 | 5 |

|  |  |  |  |  |  |  |  |
| --- | --- | --- | --- | --- | --- | --- | --- |
| **Adjectival assessment:**<br>Overall, I would rate the user-friendliness of HBM as.... | Worst imaginable | Awful | Poor | Okay | Good | Excellent | Best imaginable |
|  | 1 | 2 | 3 | 4 | 5 | 6 | 7 |

## Appendix 4

Characteristics of the accuracy study (HBM vs Philip Avalon CTG) participants who had results outside the 95% limits of agreement (>2 bpm difference). Each row shows data for one participant. No participants had more than 2 readings with >2 bpm difference. There was no association between gestation, placental position, or BMI and an excess difference in FHR (bpm).

| Subject | BMI<br>(kg/m<sup>2</sup>) | Placenta | Gestation<br>(wk) | The difference in FHR (bpm) |  |  |  |  |
| --- | --- | --- | --- | --- | --- | --- | --- | --- |
|  |  |  |  | Time 1 | Time 2 | Time 3 | Time 4 | Time 5 |
| 1 | 27.2 | posterior | 32 | 3 |  | 3 |  |  |
| 2 | 28.3 | anterior | 39 | 3 | 3 |  |  |  |
| 3 | 35.9 | anterior | 37 |  |  |  | 5 |  |
| 4 | 28.5 | posterior | 41 |  |  |  |  | 3 |
| 5 | 42.1 | lateral | 37 |  |  | 5 |  |  |
| 6 | 25.3 | anterior | 37 |  | 3 | 3 |  |  |
| 7 | 28.2 | posterior | 37 |  |  | 5 | 3 |  |

